# Computerized Cognitive Training in Cognitively Healthy Older Adults: A Systematic Review and Network Meta-Analysis

**DOI:** 10.1101/2020.10.07.20208306

**Authors:** Amit Lampit, Hanna Malmberg Gavelin, Julieta Sabates, Nathalie H Launder, Harry Hallock, Carsten Finke, Stephan Krohn, Geeske Peeters

## Abstract

**Background:** Computerized cognitive training (CCT) is a broad category of drill-and-practice interventions aims to maintain cognitive performance in older adults. Despite a supportive evidence base for general efficacy, it is unclear what types of CCT are most likely to be beneficial and what intervention design factors are essential for clinical implementation.

**Methods:** We searched MEDLINE, Embase, and PsycINFO to August 2019 for randomized controlled trials (RCTs) of any type of CCT in cognitively healthy older adults. Risk of bias within studies was assessed using the Cochrane Risk of Bias 2 tool. The primary outcome was change in overall cognitive performance between CCT and control groups. Secondary outcomes were individual cognitive domains. A series of meta-regressions were performed to estimates associations between key design factors and overall efficacy using robust variance estimation models. Network meta-analysis was used to compare the main approaches to CCT against passive or common active control conditions.

**Results:** Ninety RCTs encompassing 7219 participants across 117 comparisons were included. The overall cognitive effect size across all trials was small (*g*=0.18, 95% CI 0.14 to 0.23) with considerable heterogeneity (τ^2^=0.074, 95% prediction interval −0.36 to 0.73), robust to small-study effect or risk of bias. Effect sizes for individual cognitive domains were small, heterogeneous and statistically significant apart from fluid intelligence and visual processing. Meta-regressions revealed significantly larger effect sizes in trials using supervised training or up to three times per week. Multidomain training was the most efficacious CCT approach against any type of control, with greater benefits in a subset of supervised training studies.

**Conclusions:** The efficacy of CCT varies substantially across designs, independent of the type of control. Multidomain supervised CCT appears to be the most efficacious approach, and should be developed to accommodate for individual needs and remote delivery settings. Future research should focus on identifying the intervention components and regimens that could attenuate aging-related cognitive decline.

## INTRODUCTION

While cognitive decline is a highly common aspect of normal aging, interventions that can support cognitive function in older adults may have far-reaching health and societal implications, including delaying or preventing insidious progress towards mild cognitive impairment and dementia.^1^ In an evidence commissioned by the National Institutes of Aging, the National Academy of Medicine^2^ defined cognitive training as one of the three highest priority areas for prevention research, along with physical activity and blood pressure management. The World Health Organization guidelines for prevention of cognitive decline and dementia^3^ was similarly supportive of cognitive training, albeit based on low-quality evidence. Yet these conclusions were drawn based on an array of cognitive training interventions compared to various control conditions, leaving no guidance on how their recommendations might be implemented.

Computerized cognitive training (CCT) is a highly common cognitive training approach, based on repeated exercise repeated and controlled practice on exercises that target specific cognitive processes. CCT can be adapted to individual needs, is inherently safe and can be delivered inexpensively at scale in various clinical and community settings. About a dozen meta-analyses have investigated the efficacy of CCT in healthy older adults, generally reporting benefits for overall cognition.^4^ The largest to date, encompassing 51 randomized controlled trials (RCTs),^5^ found that efficacy could be moderated by delivery settings and session frequency but did not find differences across types of CCT and control conditions. Regardless, legitimate concerns regarding the overall quality of evidence and variability of methods in the field as well as misleading marketing practices by the “brain training” industry have driven skepticism towards CCT.^6^ Lack of clarity regarding which CCT approaches might be beneficial are therefore a clear impediment towards translating the recommendations into practice. Thus, we aimed to update and extend the findings of our previous systematic review of the field,^4^ with a particular focus on comparing the main CCT methods to the most common control conditions.

## METHODS

We followed the Preferred Reporting Items for Systematic Reviews and Meta-analyses (PRISMA) statement^7,8^ and prospectively registered the protocol PROSPERO (<u>CRD42018114891</u>). Eligibility criteria and search strategy follow our previous systematic review of the same topic.^5^

### Eligibility criteria

We considered randomized trials comparing change from baseline to post-training in one or more cognitive measure between CCT and control conditions in cognitively healthy older adults. CCT was defined as ≥4 h of practice on standardized computerized tasks or video games with clear cognitive rationale, administered on personal computers, mobile devices, or gaming consoles. Eligible controls included wait lists, alternative cognitive activities (e.g., psychoeducation) or sham conditions (e.g., low-level practice). Combinations of CCT with other interventions (e.g., physical exercise) were included if controls received the same adjacent intervention. When combined interventions were compared to passive control, trials were included if CCT comprised at least 50% of intervention time. Outcome measures that closely resembled one of more of the trained tasks were excluded.

### Information sources and study selection

We searched MEDLINE, Embase, and PsycINFO using the search terms “cognitive training” OR “brain training” OR “memory training” OR “attention training” OR “reasoning training” OR “computerized training” OR “computer training” OR “video game” OR “computer game”. No search or language limits were applied. The first search was done from inception July 2014.^5^ Search updates were applied on November 2015, February 2018 and August 2019. In each update, two or more independent reviewers performed abstract screening and assessment of full-text articles against the inclusion criteria. A senior reviewer [AL, HMG or GP] was responsible for consolidation of eligibility assessments and resolution of disagreements among reviewers. The final set of included studies was reviewed and approved by AL.

### Data items and coding

Since CCT studies typically report multiple outcome measures, all eligible measures were collected. Efficacy data were collected as mean and standard deviation (SD) for each group at each time point, or assessed using measures of change (e.g., pre-post mean and SD of change within groups). We contacted authors when reports provided insufficient data to calculate an effect size or when data for certain outcome measures were not reported. In multi-arm studies, all eligible arms were included (for a list of included arms from each study, see Table 1). Definitions of contrasts for the NMA occasionally differed from the pairwise meta-analyses, especially in multi-arm trials to reflect all available comparisons. Coding CCT and active control conditions into specific types was done based on the content of the intervention. Coding of outcome measures into specific cognitive domains was done based on the Cattell-Horn-Carroll-Miyake framework.^9^

**Table 1:**
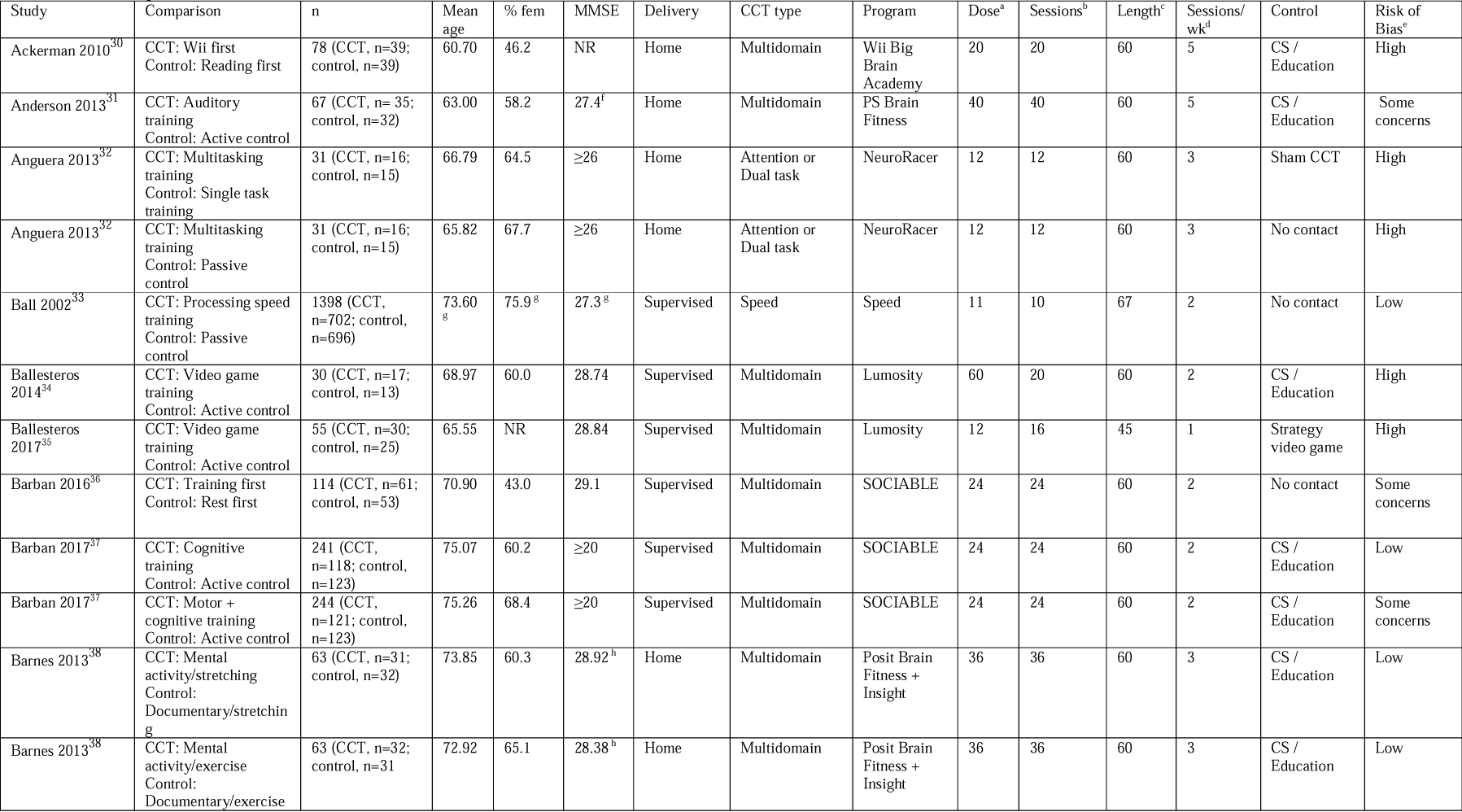
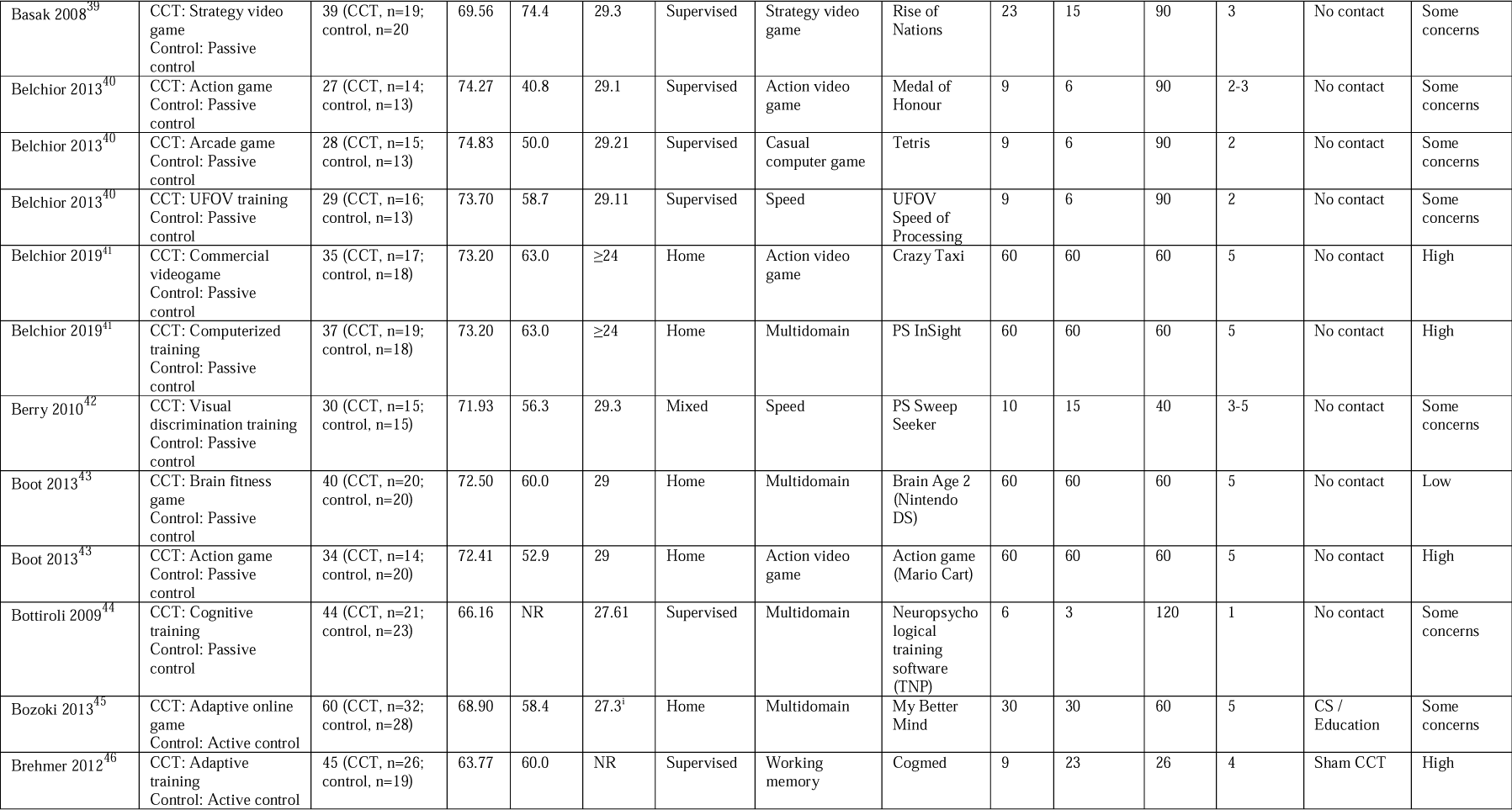
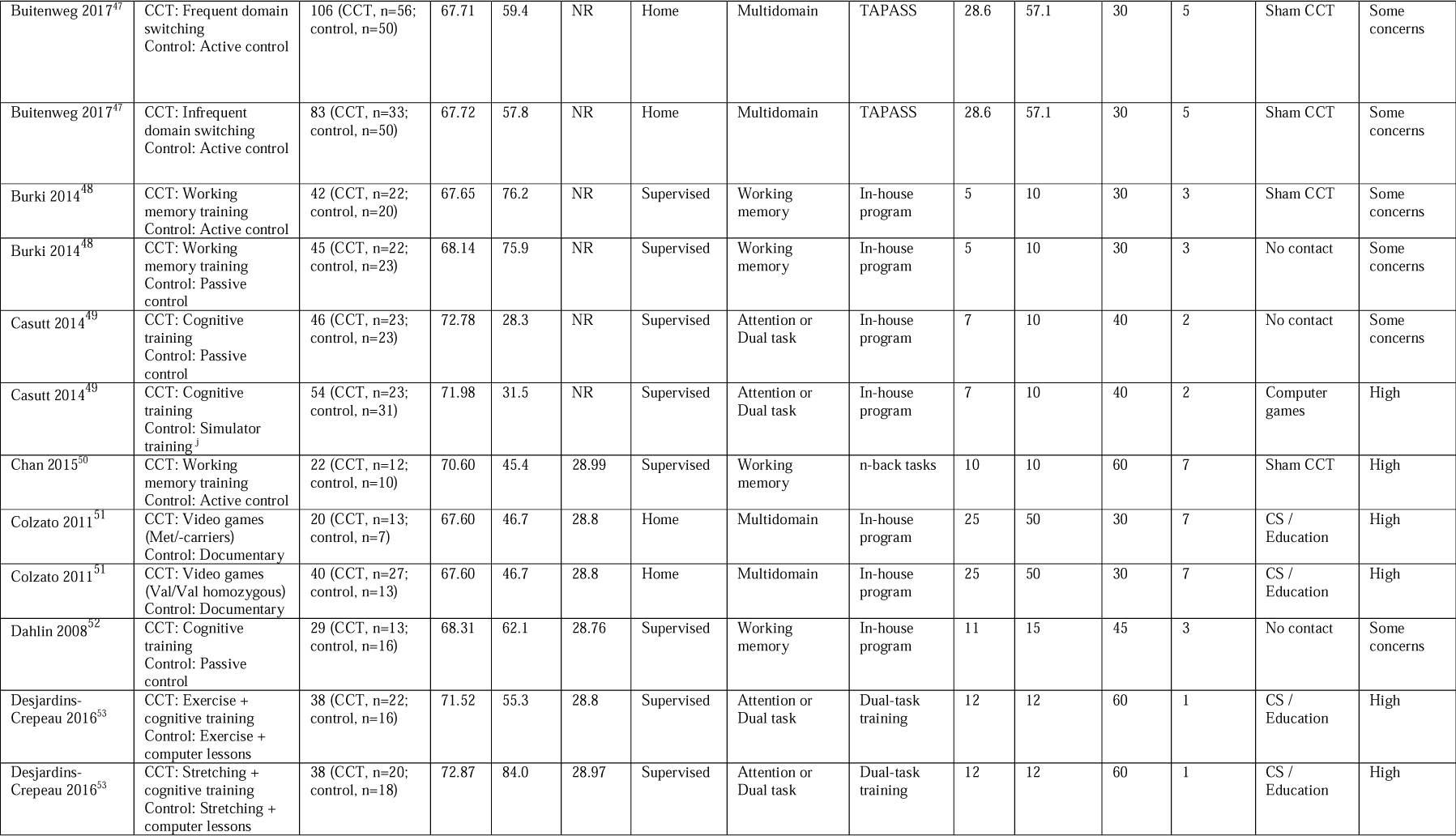
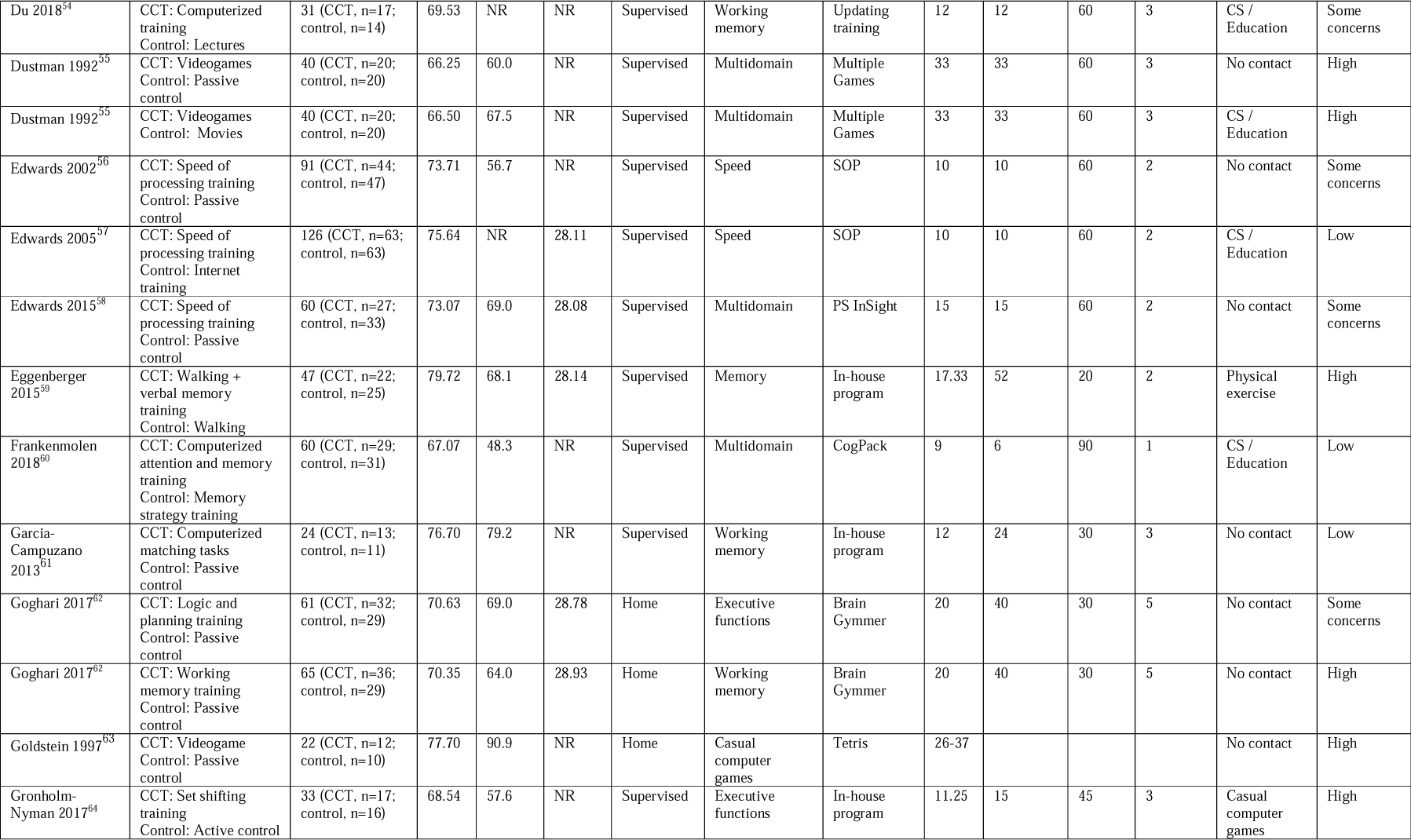
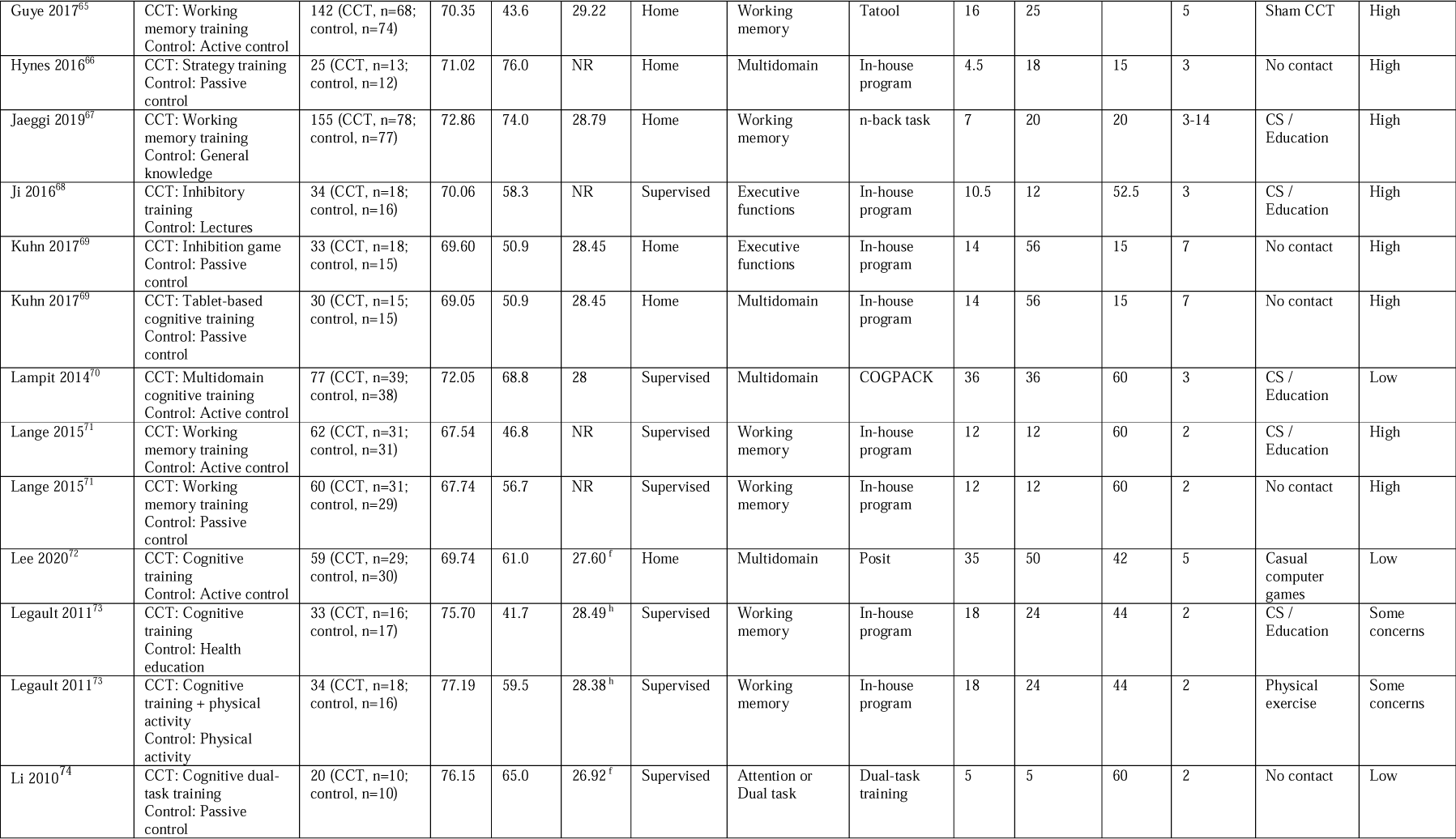
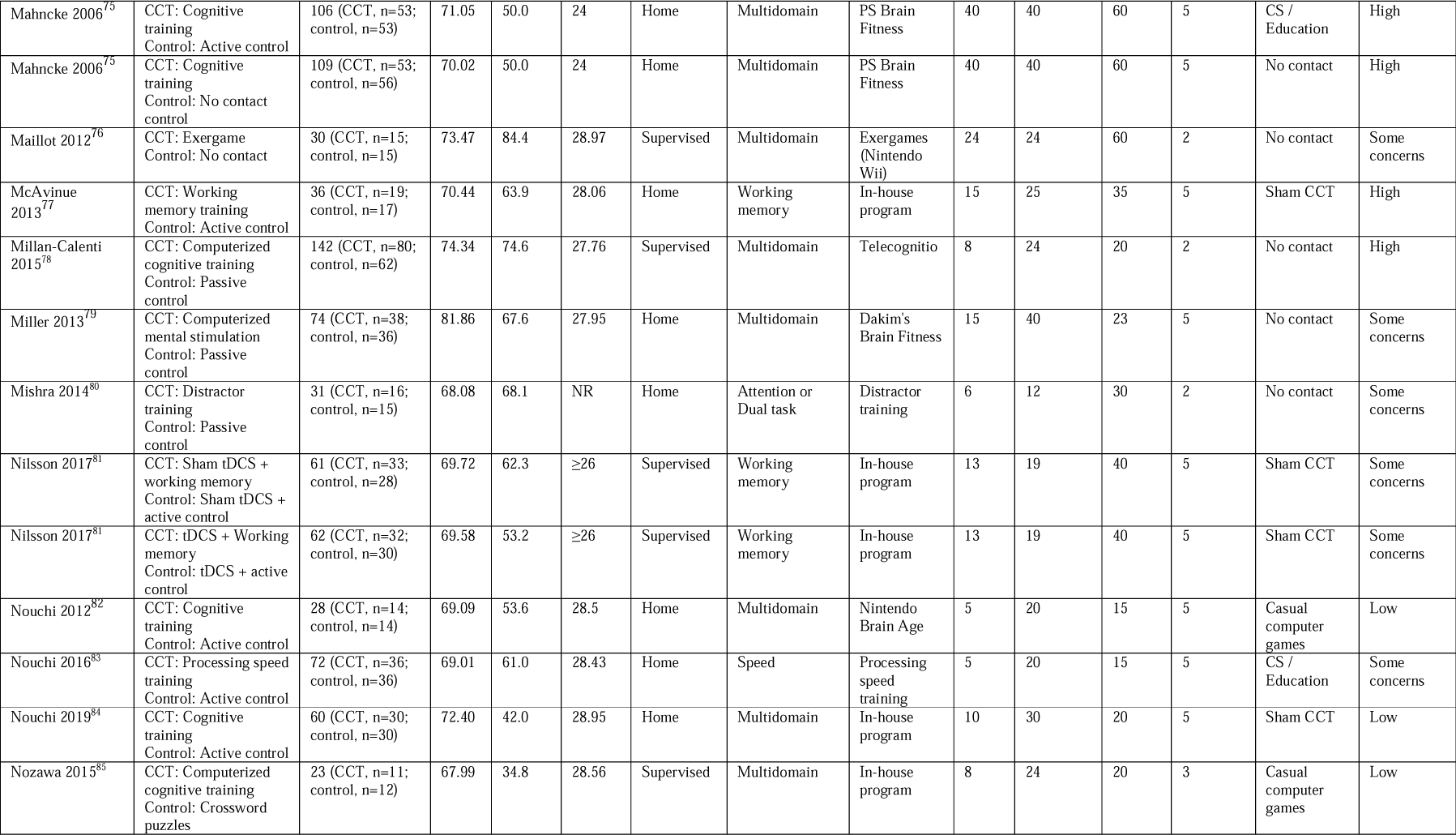
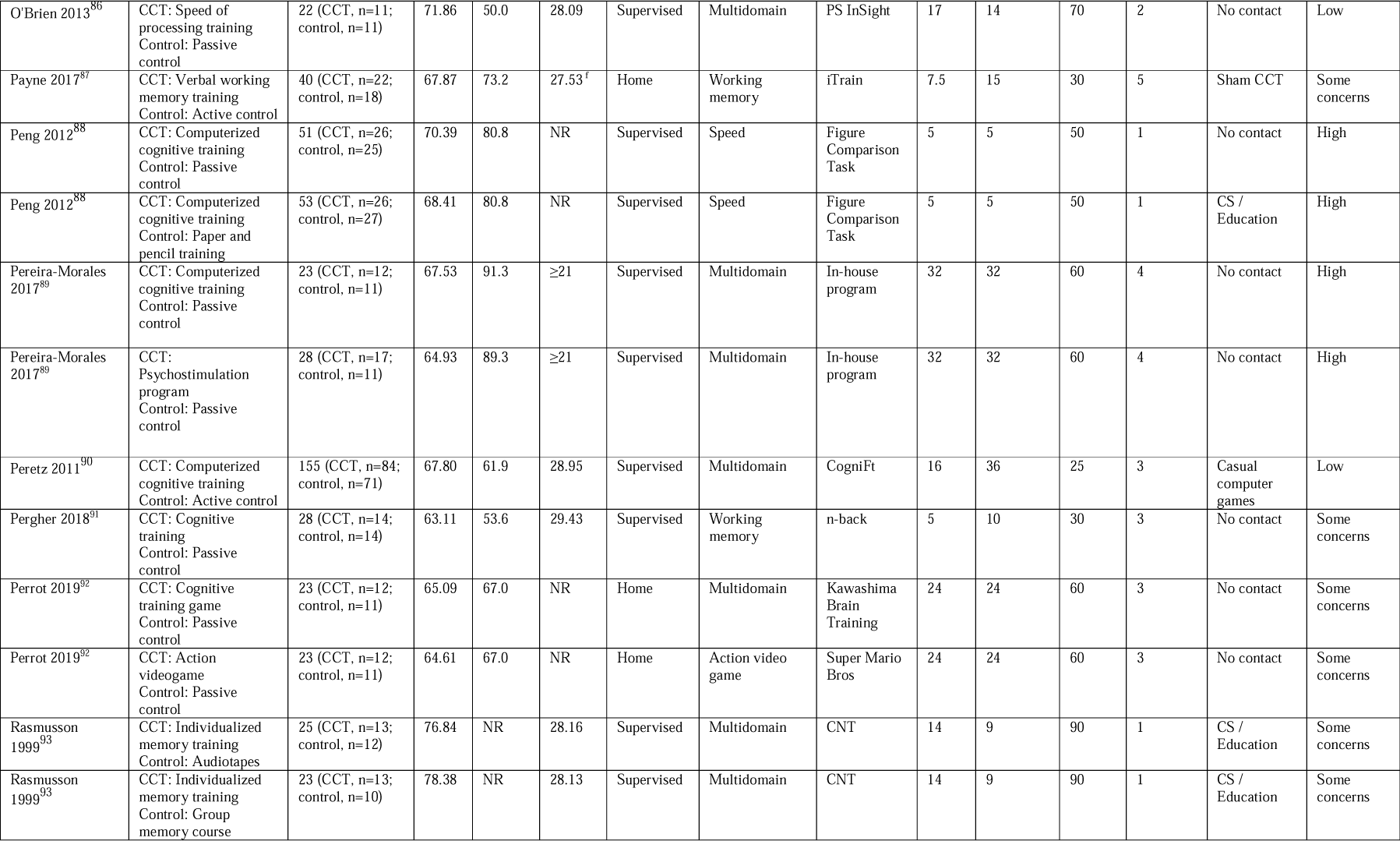
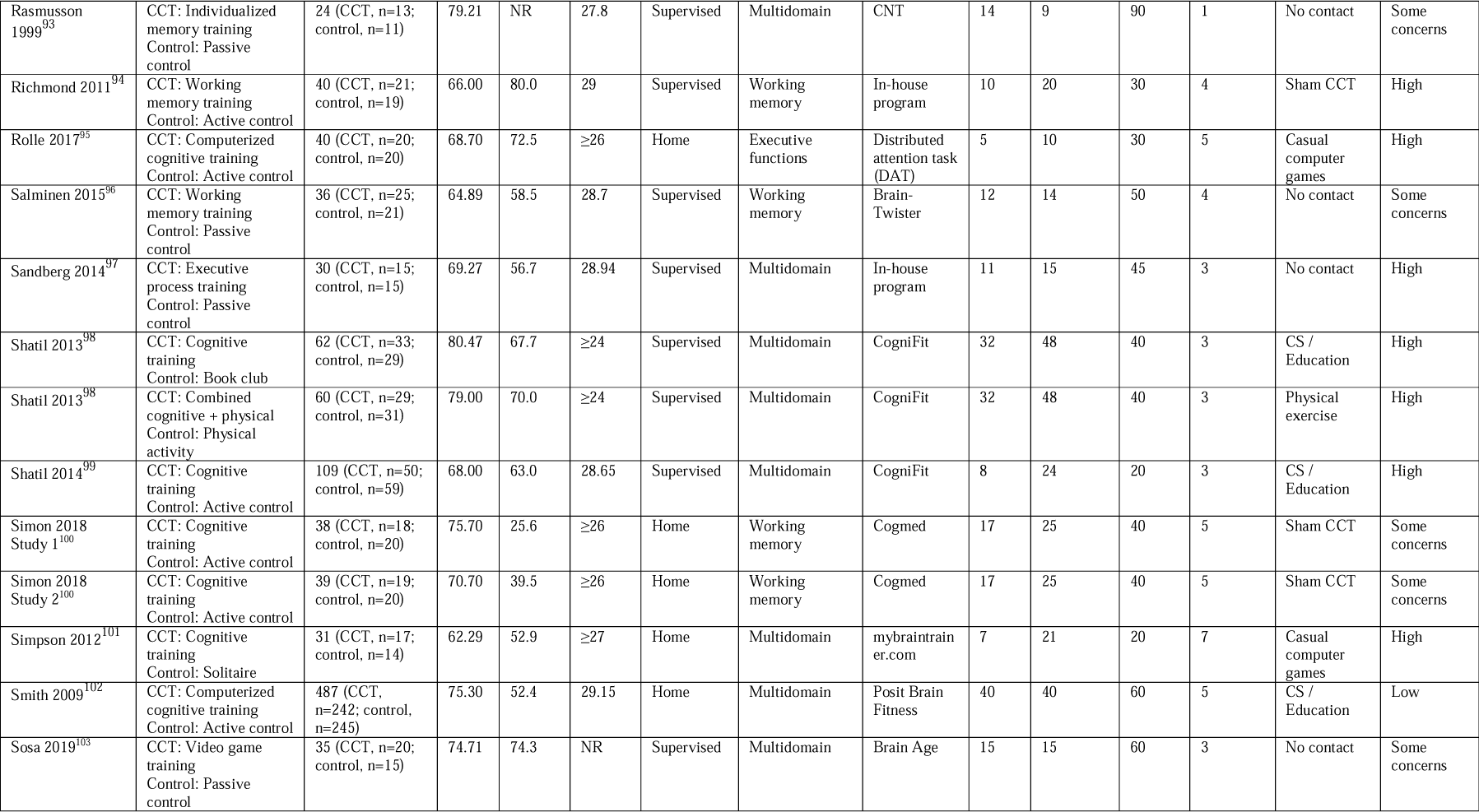
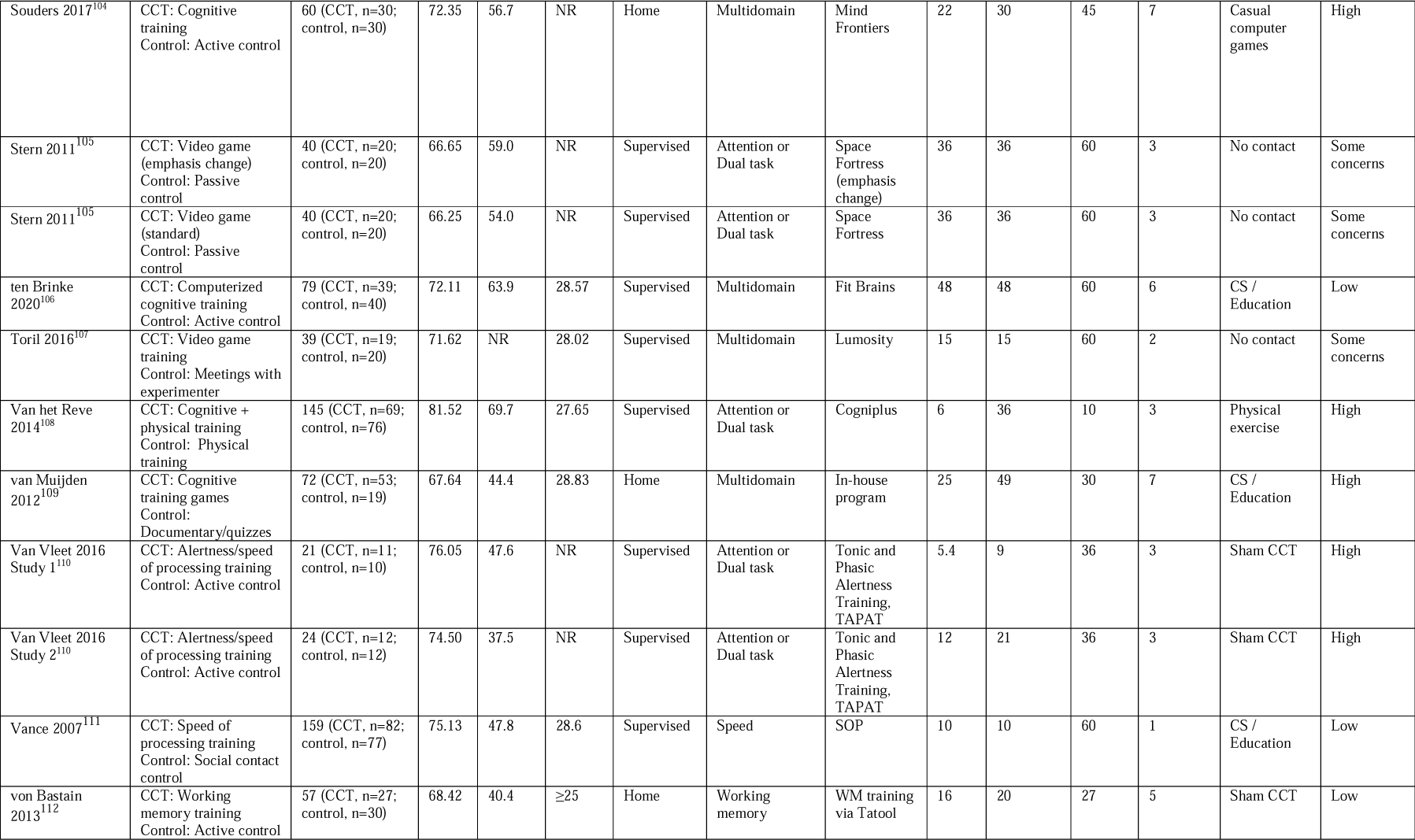
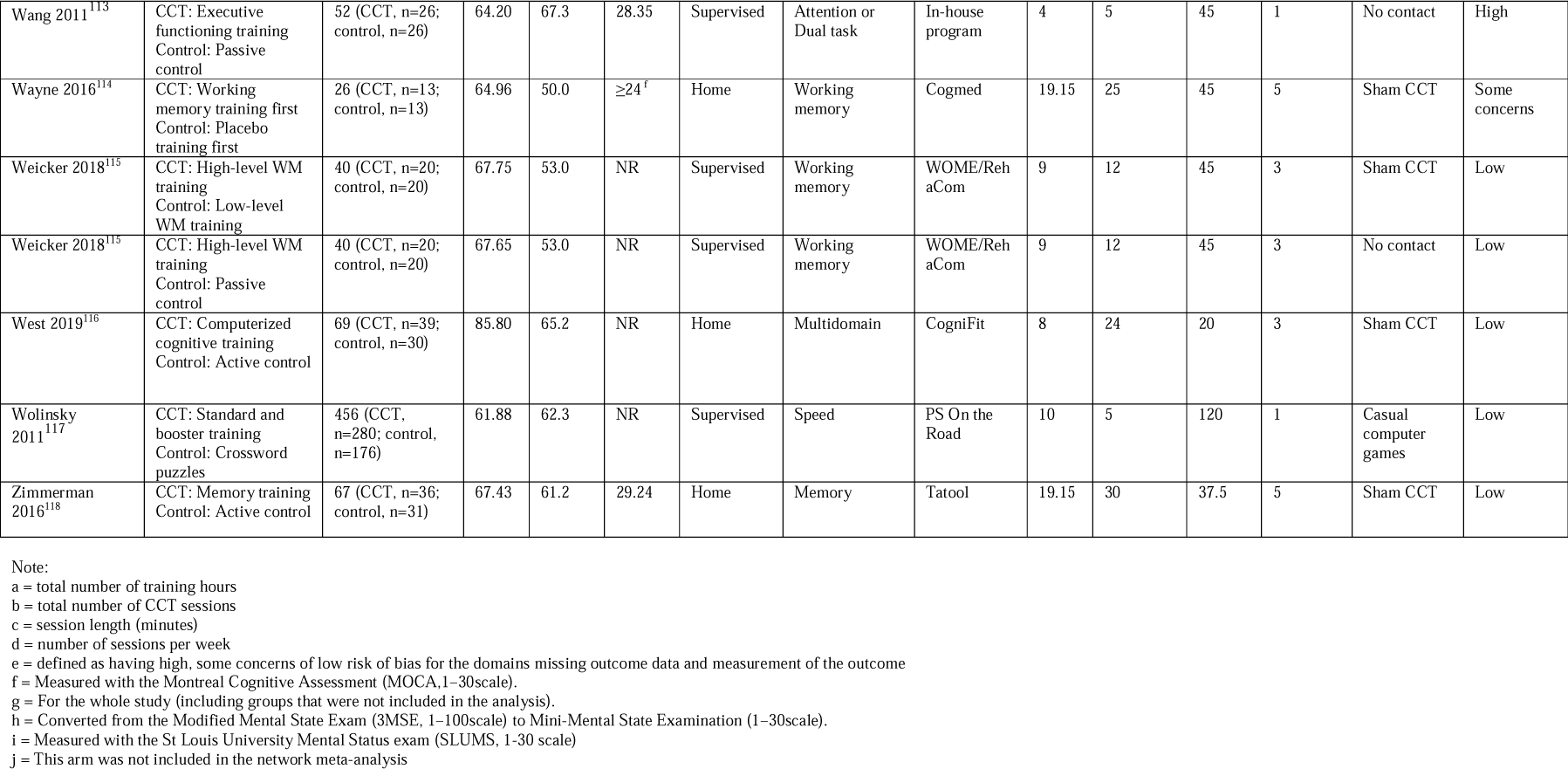
Study characteristics.

### Risk of bias within studies

We used the 2019 Cochrane Risk of Bias 2 tool^10^ (RoB2) to assess risk of bias across five domains (randomization process, deviations from intended interventions, missing outcome data, measurement of the outcome, selection of the reported result). In addition, each study received an overall RoB assessment of high, low or some concerns. In contrast to the original RoB2 macros, studies that did not report assessor blinding or intention-to-treat results were coded as high risk of bias regardless of other RoB2 items.

### Statistical analysis

Analyses were conducted using the R packages robumeta,^11^ clubSandwich^12^ and netmeta.^13^ The primary outcome was overall cognition, defined as a composite of all eligible outcomes reported in each trial.^5^ Secondary outcomes were individual cognitive domains. Between-group differences in each outcome measure were converted to standardized mean differences and calculated as Hedges’ g with 95% CI. Pairwise analyses were performed using robust variation estimation^14^ (RVE) with robumeta, based on correlational dependence model with r=0.8. Heterogeneity across studies was quantified using τ^2^ and expressed as a proportion of overall observed variance using the I^2^ statistic.^15^ Prediction intervals were calculated to assess the dispersion of true effects across studies.^16^ RVE meta-regressions based on prespecified categorical moderators were performed using robumeta and formally tested for between-group differences based on F-statistic using clubSandwich. Small-study effect for the primary outcome was investigated by visually inspecting funnel plots of effect size vs standard error and formally tested using the Egger’s test as a meta-regression in RVE.^17,18^ Second, random-effects network meta-analysis of the primary and secondary outcomes was performed using a frequentist framework using netmeta. Network geometry was summarized in a network graph and league tables were created to display the relative effect sizes of all available comparisons. Ranking of treatments were estimated using P-scores, representing the extent of certainty that an intervention is more effective than another intervention^19^. Higher P-scores represent higher likelihood of a certain intervention to be the more effective. To examine the transitivity assumptions, we created a table summarizing potential effect modifiers (design characteristics and risk of bias) to explore whether these were similarly distributed across the different comparisons. Sensitivity analysis of the primary outcome were conducted for subsets of supervised and home-based training studies.

## RESULTS

After accounting for duplicates within and across searches, we screened 14,361 unique titles, of which 762 full-text articles were assessed against the inclusion criteria, resulting in 90 eligible RCTs encompassing 7219 participants (**Fig 1**). Fourteen RCTs were identified from manual searches and four potentially eligible RCTs were excluded because the reports did not provide sufficient data for analysis and authors did not provide data following our requests.

**Figure 1:**
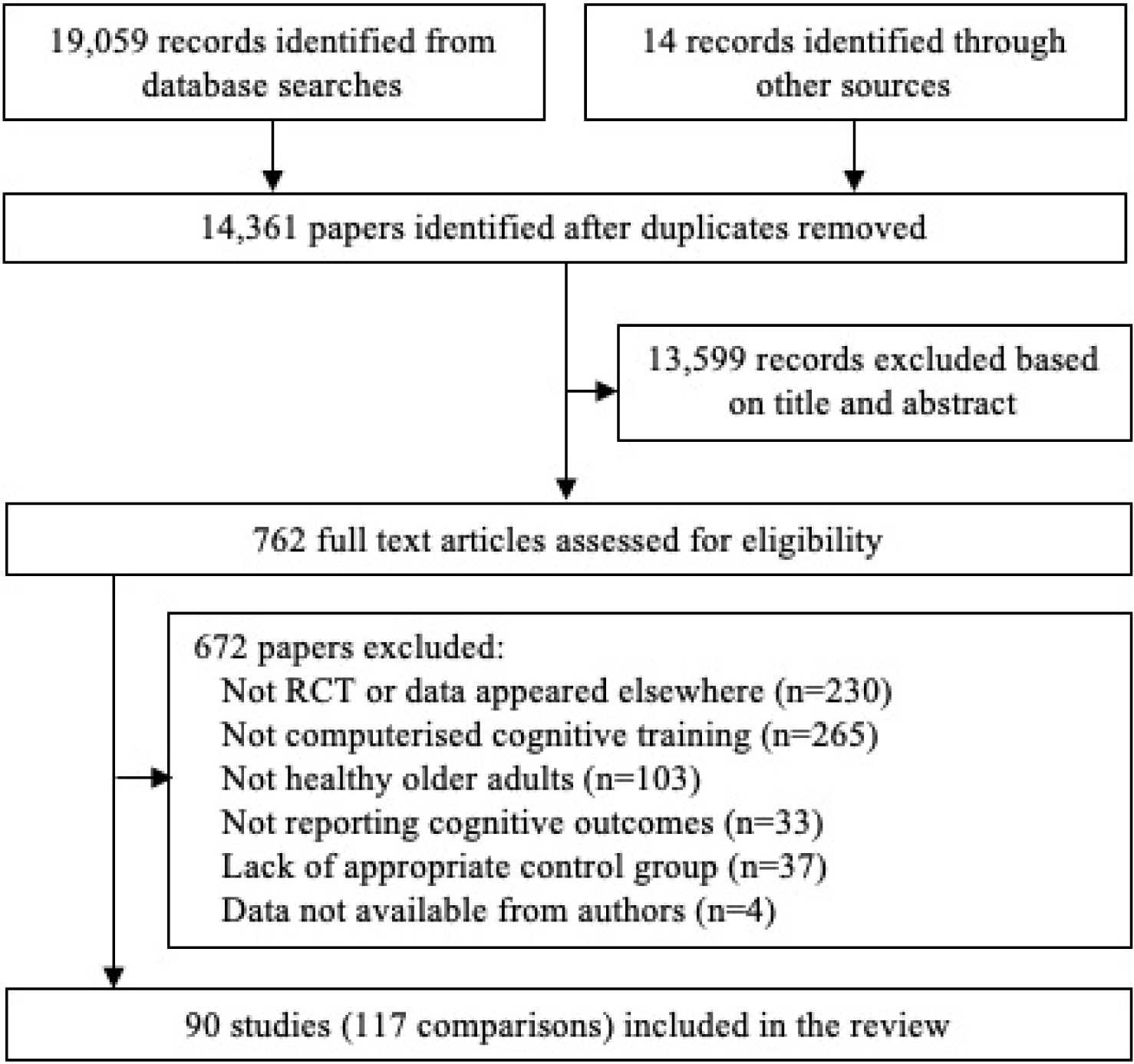
PRISMA flowchart.

### Characteristics of included studies

Key study characteristics are reported in **Table 1**. The 90 RCTs included 117 eligible CCT and control arms. The most common type of CCT was multidomain training (n=36 RCTs), followed by working memory training (n=21) and attention/dual task CCT (n=10). Fifty-nine trials (66%) compared CCT to at least one active control condition, of which 11 included an additional passive control group, and 7 trials included more than one CCT arm (**Figure 2**). Overall risk of bias was assessed as low in 21 trials, high in 36, and 29 had some concerns (**Table 1**).

**Figure 2:**
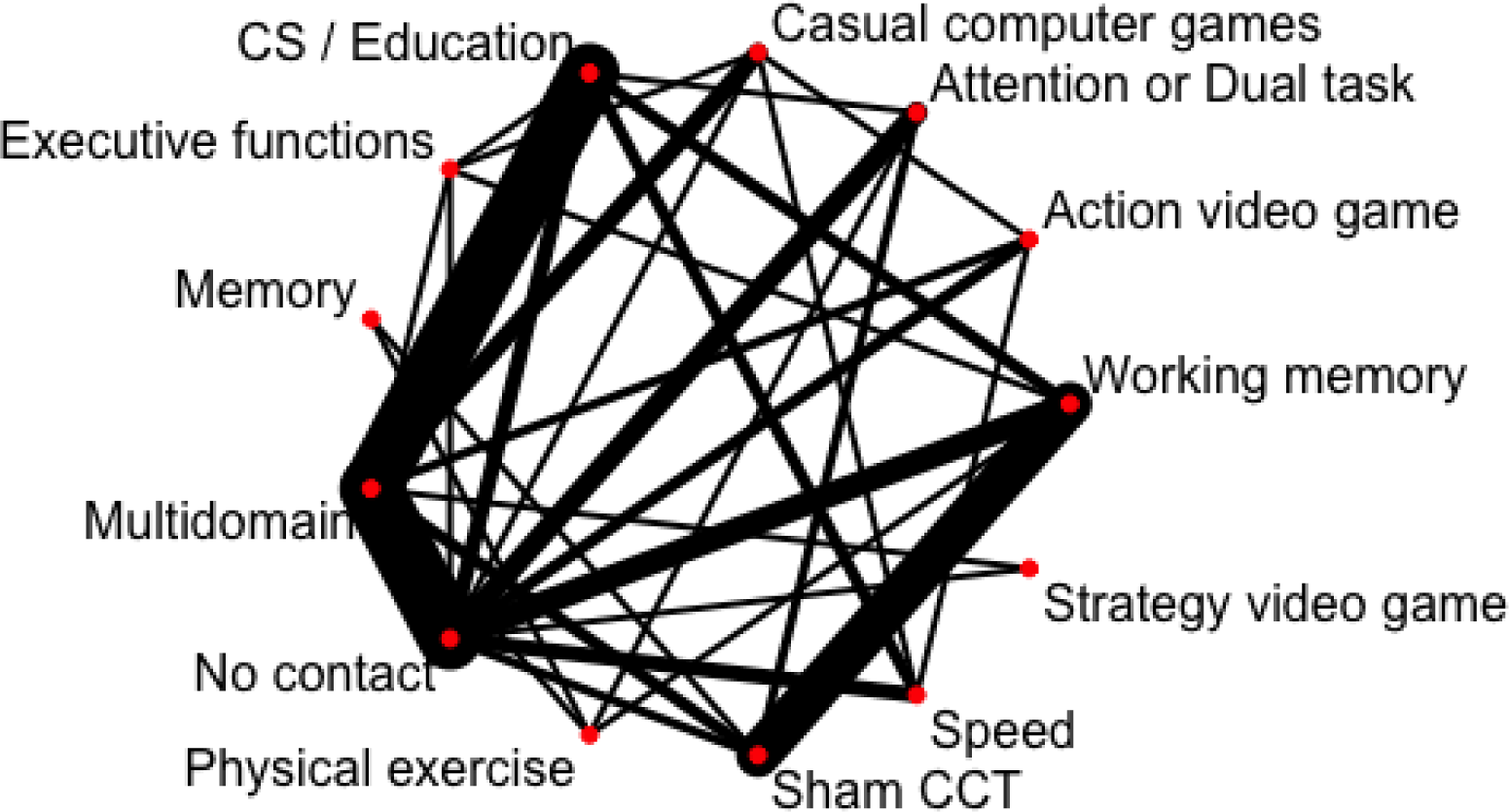
Network structure.

### Primary outcome: Overall cognition

The pooled overall effect size across all 90 RCTs (1211 effect sizes, median 10 effect sizes per study) was small (*g*=0.18, 95% CI 0.14 to 0.23) with considerable heterogeneity (τ^2^=0.074, I^2^=58%). The 95% prediction interval indicated high variability in overall effect sizes across settings (−0.36 to 0.73). There was no evidence for small-study effect (β=0.35, one-tailed p=0.117, Figure 3). There was no evidence for difference across levels of risk of bias (F_2,64.8_=0.391, p=0.678).

**Figure 3:**
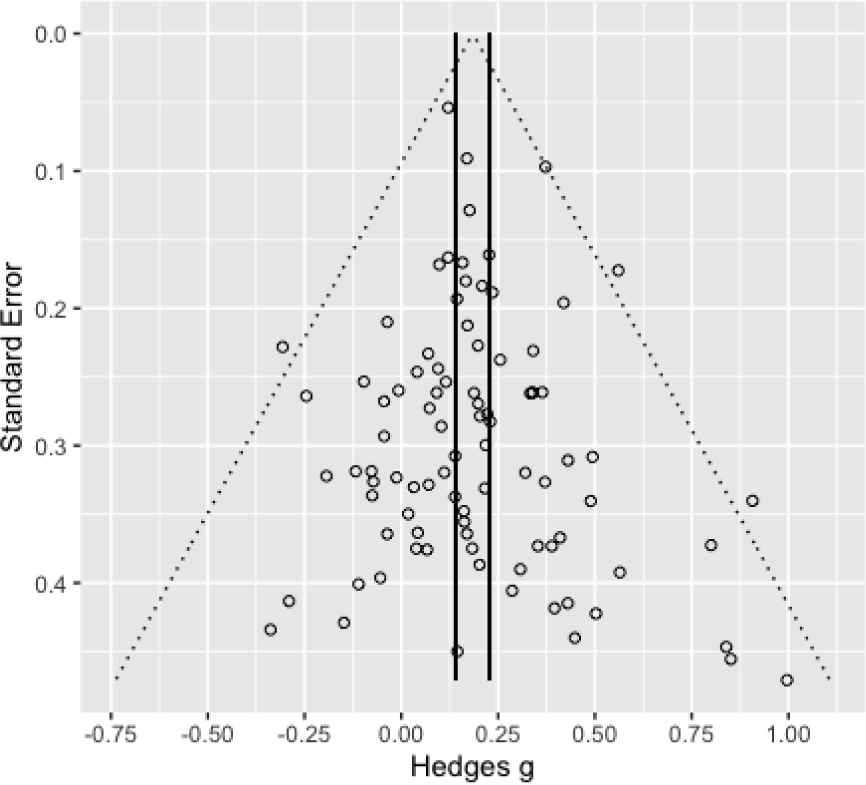
Funnel plot (primary outcome)

### Meta-regressions

Results of meta-regressions for the primary outcome are provided in **Table 2**. The pooled effect size was significant larger for supervised vs home-based training (F_1,68.5_=5.8, p=0.019) and for training 1-3 times per week vs more frequent regimens (F_1,71.2_=4.9, p=0.029). Session length, treatment duration and total hours of training were not associated with overall cognitive effect size. Compared to studies that used supervised training, home-based training studies tended to provide more frequent (t_74.8_=8.82, p<0.001) and shorter sessions (t_87.5_=-3.67, p<0.001), as well as more hours of training (t_55.4_=2.19, p=0.032). Multiple meta-regressions did not find interactions between delivery mode and any dosing factor.

**Table 2:** Tests of moderators (primary outcome)

| Moderator | No. of studies (effect sizes) | Summary effect |  |  |  | Test of moderation |  |  |
| --- | --- | --- | --- | --- | --- | --- | --- | --- |
|  |  | Hedges' *g* (95% CI) | *t* | df | *p* | *F* | df | *p* |
| **Delivery** |  |  |  |  |  | 5.8 | 1,68.5 | 0.018 |
| Supervised | 54 (731) | 0.23 (0.17 to 0.28) | 7.98 | 47.3 | <0.001 |  |  |  |
| Home-based | 36 (470) | 0.12 (0.05 to 0.19) | 3.55 | 32.1 | 0.001 |  |  |  |
| **Frequency** |  |  |  |  |  | 4.93 | 1,72.2 | 0.029 |
| 1-3 per week | 53 (706) | 0.23 (0.17 to 0.28) | 7.58 | 46.2 | <0.001 |  |  |  |
| 4-7 per week | 37 (495) | 0.13 (0.06 to 0.19) | 3.91 | 33.2 | <0.001 |  |  |  |
| **Session length** |  |  |  |  |  | 0.57 | 2,54 | 0.568 |
| 1-30 min | 28 (312) | 0.15 (0.06 to 0.24) | 3.48 | 24.9 | 0.002 |  |  |  |
| >30 – <60 min | 23 (373) | 0.18 (0.11 to 0.26) | 5.19 | 21.9 | <0.001 |  |  |  |
| ≥60 min | 39 (516) | 0.21 (0.14 to 0.29) | 5.65 | 32.5 | <0.001 |  |  |  |
| **Training duration** |  |  |  |  |  | 0.48 | 2,49.9 | 0.62 |
| ≤9 hours | 25 (242) | 0.21 (0.11 to 0.31) | 4.28 | 22.0 | <0.001 |  |  |  |
| >9 – 20 | 42 (591) | 0.16 (0.09 to 0.23) | 4.94 | 37.1 | <0.001 |  |  |  |
| >20 hours | 23 (368) | 0.20 (0.11 to 0.29) | 4.87 | 20.0 | <0.001 |  |  |  |
| **Risk of bias** |  |  |  |  |  | HTZ<br>1.38 | 4.95 | 0.33 |
| Low | 21 (273) | 0.15 (0.09 to 0.22) | 4.71 | 21.2 | <0.001 |  |  |  |
| Some concerns | 29 (421) | 0.19 (0.11 to 0.26) | 5.20 | 21.2 | <0.001 |  |  |  |
| High | 36 (507) | 0.19 (0.13 to 0.26) | 5.86 | 45.1 | <0.001 |  |  |  |

### Secondary outcomes: Individual cognitive domains

Meta-analyses of individual cognitive domains are provided in **Table 3**. Effect sizes across the six domains were generally small and heterogeneous. There was no evidence for benefit on fluid intelligence, and the pooled estimate for visual processing did not reach statistical significance.

**Table 3:** Analyses of individual cognitive domains.

| Broad CHCM domain | No. of studies (effect sizes) | Hedges' *g* | *t* | df | *p* | $\tau^2$ | $I^2$ |
| --- | --- | --- | --- | --- | --- | --- | --- |
| Executive functions | 59 (535) | 0.17 (0.11 to 0.23) | 5.82 | 50.2 | <0.001 | 0.036 | 32.7 |
| Fluid intelligence | 33 (95) | 0.08 (-0.04 to 0.20) | 1.35 | 28.1 | 0.187 | 0.051 | 48.3 |
| Long-term memory and retrieval | 45 (156) | 0.16 (0.06 to 0.26) | 3.29 | 37.5 | 0.002 | 0.052 | 51.7 |
| Processing speed | 62 (235) | 0.21 (0.12 to 0.31) | 4.37 | 59.6 | <0.001 | 0.231 | 80.6 |
| General short-term memory | 63 (261) | 0.17 (0.11 to 0.24) | 5.49 | 53.1 | <0.001 | 0.038 | 35.9 |
| Visual processing | 28 (88) | 0.12 (-0.01 to 0.24) | 2 | 25.1 | 0.057 | 0.072 | 45.1 |

### Network meta-analysis: Primary outcome

The 90 RCTs provided 131 pairwise comparisons across 13 CCT or control conditions, resulting in a well-connected network structure (**Figure 2**). Direct evidence was available for 32 comparisons, most notably multidomain vs no contact (20 RCTs), multidomain vs CS/Education (19 RCTs) and working memory training vs sham (14 RCTs). There was evidence for inconsistency for four comparisons; the direct effect size was larger than the indirect estimate for multidomain vs no contact and speed vs casual computer games, and smaller for speed vs no contact (**Table 4**).

**Table 4:** Network meta-analysis results (primary outcome)

|  |  |  |  |  |  |  |  |  |  |  |  |  |
| --- | --- | --- | --- | --- | --- | --- | --- | --- | --- | --- | --- | --- |
| Multidomain |  |  |  | 0.07<br>[-0.41, 0.56] | 0.00<br>[-0.37, 0.38] | 0.07<br>[-0.57, 0.71] |  | **0.15**<br>**[0.07, 0.24]** | 0.26<br>[-0.21, 0.72] | 0.08<br>[-0.14, 0.30] | **0.30**<br>**[0.18, 0.42]** | 0.18<br>[-0.02, 0.38] |
| 0.03<br>[-0.08, 0.13] | Speed |  |  |  | 0.24<br>[-0.46, 0.95] |  |  | **0.20**<br>**[0.02, 0.38]** |  |  | **0.14**<br>**[0.05, 0.23]** | **0.35**<br>**[0.17, 0.54]** |
| 0.04<br>[-0.14, 0.22] | 0.01<br>[-0.18, 0.20] | Attention or<br>Dual task |  |  |  |  |  | 0.07<br>[-0.34, 0.48] | 0.16<br>[-0.14, 0.45] | **0.50**<br>**[0.07, 0.92]** | 0.11<br>[-0.12, 0.33] |  |
| 0.05<br>[-0.08, 0.19] | 0.02<br>[-0.12, 0.17] | 0.01<br>[-0.19, 0.21] | Working<br>memory |  |  | 0.03<br>[-0.40, 0.47] |  | 0.12<br>[-0.10, 0.33] | 0.10<br>[-0.51, 0.72] | **0.14**<br>**[0.00, 0.27]** | 0.17<br>[-0.03, 0.37] |  |
| 0.04<br>[-0.33, 0.42] | 0.01<br>[-0.37, 0.40] | 0.00<br>[-0.41, 0.41] | -0.01<br>[-0.40, 0.38] | Strategy<br>video game |  |  |  |  |  |  | 0.22<br>[-0.37, 0.80] |  |
| 0.07<br>[-0.23, 0.36] | 0.04<br>[-0.26, 0.34] | 0.03<br>[-0.31, 0.36] | 0.02<br>[-0.30, 0.33] | 0.03<br>[-0.45, 0.50] | Action video<br>game |  |  |  |  |  | 0.20<br>[-0.13, 0.54] | -0.21<br>[-0.89, 0.48] |
| 0.16<br>[-0.09, 0.40] | 0.13<br>[-0.12, 0.38] | 0.12<br>[-0.18, 0.41] | 0.10<br>[-0.15, 0.36] | 0.11<br>[-0.33, 0.56] | 0.09<br>[-0.28, 0.46] | Executive<br>functions |  | 0.16<br>[-0.45, 0.77] |  |  | -0.02<br>[-0.39, 0.35] | 0.01<br>[-0.41, 0.43] |
| 0.18<br>[-0.18, 0.55] | 0.16<br>[-0.21, 0.53] | 0.14<br>[-0.23, 0.52] | 0.13<br>[-0.23, 0.49] | 0.14<br>[-0.38, 0.66] | 0.12<br>[-0.35, 0.58] | 0.03<br>[-0.40, 0.46] | Memory |  | -0.04<br>[-0.57, 0.48] | 0.04<br>[-0.40, 0.48] |  |  |
| **0.17**<br>**[0.09, 0.24]** | **0.14**<br>**[0.03, 0.25]** | 0.13<br>[-0.06, 0.31] | 0.12<br>[-0.02, 0.25] | 0.12<br>[-0.26, 0.50] | 0.10<br>[-0.20, 0.40] | 0.01<br>[-0.23, 0.26] | -0.02<br>[-0.38, 0.35] | CS /<br>Education |  |  | 0.04<br>[-0.18, 0.26] |  |
| 0.20<br>[-0.05, 0.44] | 0.17<br>[-0.09, 0.42] | 0.16<br>[-0.08, 0.39] | 0.14<br>[-0.11, 0.40] | 0.15<br>[-0.29, 0.60] | 0.13<br>[-0.25, 0.50] | 0.04<br>[-0.30, 0.37] | 0.01<br>[-0.36, 0.38] | 0.03<br>[-0.22, 0.28] | Physical<br>exercise |  |  |  |
| **0.18**<br>**[0.04, 0.33]** | **0.16**<br>**[0.00, 0.31]** | 0.14<br>[-0.06, 0.35] | **0.13**<br>**[0.01, 0.25]** | 0.14<br>[-0.25, 0.54] | 0.12<br>[-0.20, 0.44] | 0.03<br>[-0.24, 0.29] | 0.00<br>[-0.35, 0.35] | 0.02<br>[-0.13, 0.16] | -0.01<br>[-0.27, 0.25] | Sham CCT | 0.05<br>[-0.28, 0.39] |  |
| **0.21**<br>**[0.12, 0.30]** | **0.19**<br>**[0.10, 0.27]** | **0.17**<br>**[0.00, 0.35]** | **0.16**<br>**[0.03, 0.29]** | 0.17<br>[-0.21, 0.55] | 0.15<br>[-0.15, 0.44] | 0.06<br>[-0.18, 0.30] | 0.03<br>[-0.34, 0.39] | 0.05<br>[-0.05, 0.14] | 0.02<br>[-0.23, 0.26] | 0.03<br>[-0.12, 0.17] | No contact | -0.45<br>[-0.99, 0.09] |
| **0.25**<br>**[0.11, 0.39]** | **0.22**<br>**[0.08, 0.36]** | 0.21<br>[-0.01, 0.43] | **0.20**<br>**[0.02, 0.38]** | 0.21<br>[-0.19, 0.60] | 0.18<br>[-0.13, 0.50] | 0.09<br>[-0.16, 0.35] | 0.07<br>[-0.32, 0.45] | 0.08<br>[-0.07, 0.23] | 0.06<br>[-0.22, 0.33] | 0.07<br>[-0.12, 0.26] | 0.04<br>[-0.10, 0.18] | Casual<br>computer<br>games |

Across all trials, multidomain training ranked highest for efficacy on overall cognition, with small and statistically significant effect sizes over and above passive control (g=0.21, 95% CI 0.12 to 0.30) and all active control conditions apart from physical exercise (Table 4). Processing speed training was ranked second with similar but slightly smaller estimates. Working memory training was better than all control conditions apart from cognitive stimulation. There was no evidence for cognitive benefit of any active control condition over and above no contact control.

When separating supervised and home-based training studies, only multidomain training was found to be more efficacious than passive control (g=0.30, 95% CI 0.18 to 0.41) and CS/Education (g=0.25, 95% CI 0.14 to 0.36). There was no evidence of benefit for any home-based condition. Finally, an RVE analysis of the 21 RCTs that used supervised multidomain CCT revealed a similar estimate (g=0.30, 95% CI 0.20 to 0.40), with about half the heterogeneity of the full model (τ^2^=0.037, I^2^=36%). Of these, 19 provided training up to 3 times per week, resulting in nearly identical estimates (g=0.30, 95% CI 0.19 to 0.41, τ^2^=0.038, I^2^=38%).

#### Secondary outcomes

Network meta-analyses ranking for individual domains are presented in Figure 4. The CCT types ranked highest and reported statistically significant benefits were multidomain (g=0.24, 95% CI 0.10 to 0.38) and working memory training (g=0.22, 95% CI 0.06 to 0.38) for executive functions, speed (g=0.36, 95% CI 0.08 to 0.65) and multidomain (g=0.26, 95% CI 0.05 to 0.46) for long-term memory and retrieval, speed (g=0.61, 95% CI 0.38 to 0.83) and multidomain (g=0.36, 95% CI 0.18 to 0.54) for processing speed, and attention/dual task (g=0.46, 95% CI 0.21 to 0.72) and multidomain (g=0.19, 95% CI 0.05 to 0.32) for general short-term memory. Analyses of fluid intelligence and visual processing did not identify statistically significant benefits for any CCT type.

**Figure 4:**
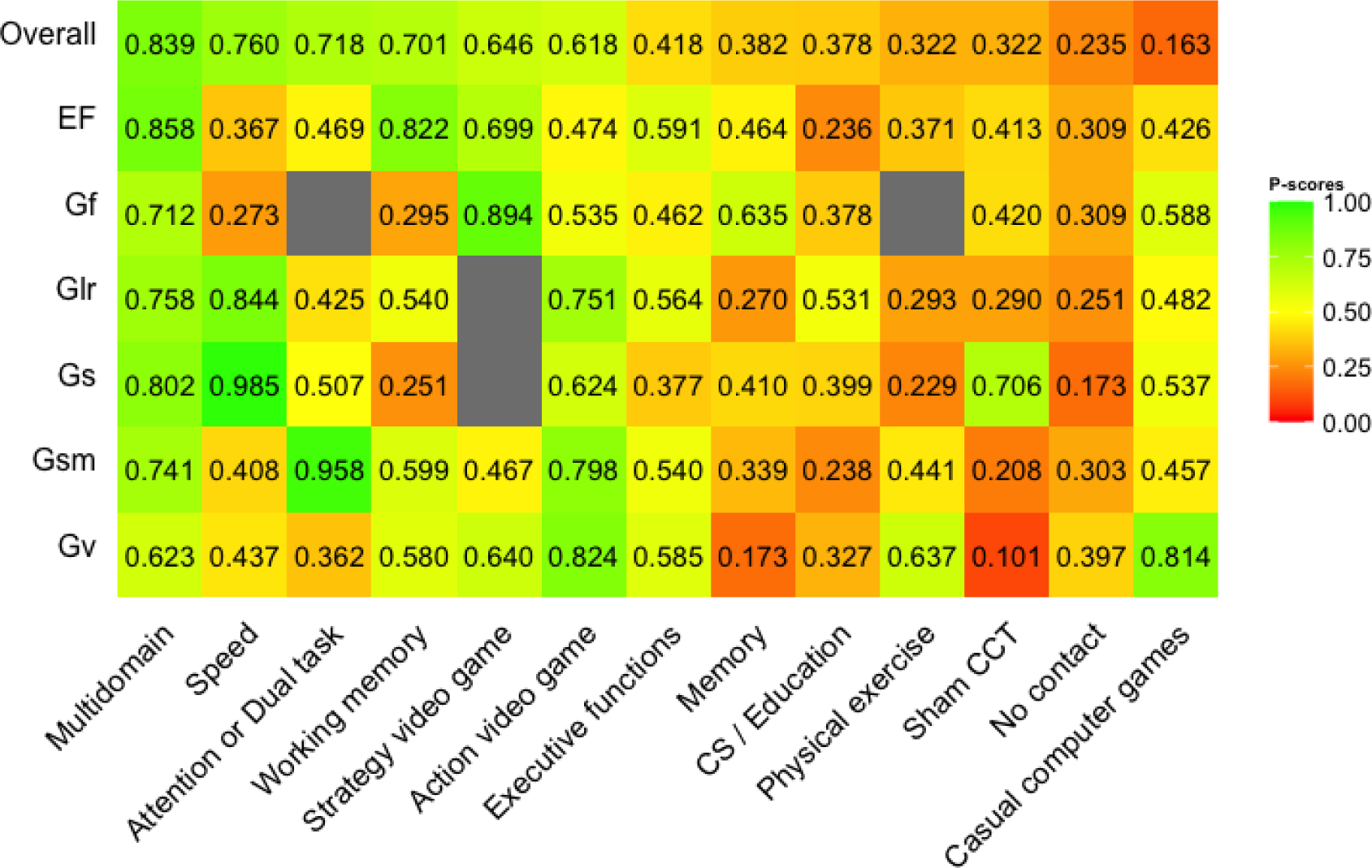
P-scores for primary and secondary outcomes.

## DISCUSSION

This multivariate and network meta-analysis of 90 RCTs has confirmed the efficacy of CCT and narrowed down the conditions in which CCT can result in cognitive benefits in healthy older adults. Our results suggest that multidomain CCT as the most sensible approach to improving global and domain-specific cognitive performance in this population, and that these effects can be further augmented by implementing supervised settings across up to three weekly sessions. However, trials that used supervised CCT were also more likely to provide less frequent sessions, and therefore it was not possible to examine whether these two factors are independent effect modifiers.

Furthermore, comparisons of common active control conditions (general cognitive stimulation, sham CCT and computer games) do not point to a benefit over and above no-contact (passive) control, suggesting that these are ineffectual not only as stand-alone interventions, but also as a means to control for non-specific (‘placebo’) effects in CCT trials apart from those associated with repeated testing.

The effect size estimates for the primary outcome as well as lack of evidence for small-study effect or association with risk of bias are consistent with our previous meta-analysis,^5^ and so are the role of supervision and session frequency as effect modifiers. The current meta-analysis included 40 additional RCTs that meet the same stringent eligibility criteria and used more efficient methods to handle dependent effect sizes in meta-regressions. Thus, these findings are likely to be robust and substantially increase the certainty in the RCT evidence for the general efficacy of CCT.

Given the wealth of RCTs, reasonable certainty in the evidence and lack of evidence to support the efficacy of active or passive control conditions, clinical equipoise assumptions in CCT trials are becoming increasingly difficult to justify. That is, the new knowledge gained from clinical trials comparing CCT programs to inert control may not necessarily be substantial enough to withhold potentially effective intervention from older participants.^20^ At the same time, the opportunity cost of testing relatively basic efficacy hypotheses (‘does CCT work?’) instead of addressing clinical implementation challenges in the field is increasing. Specifically, in order to test the effectiveness of CCT as a means to prevent cognitive decline at scale, research that focuses on maintaining engagement over time, providing remote supervision akin to that of center-based training and personalizing CCT, among other research priorities. These would require larger and longer studies that those typically conducted in the field but allow investigations of novel approaches compared to existing CCT programs.

Our network meta-analysis highlights the importance of multidomain training as the CCT approach most likely to be beneficial. Several meta-analyses of single domain training have shown that training gains tend to be most pronounced within the domains targeted by the program.^21,22^ Lack of robust evidence for gains in untrained domains is often cited as a limitation of CCT but in fact this is simply the reality of most interventions targeting a specific physiological or mental process. From a clinical implementation perspective, targeted CCT may be clinically relevant for rehabilitation of specific cognitive deficits, such as a recent FDA-approved attention training program for children with attention-deficit/hyperactivity disorder.^23^ However, given age-related cognitive decline affects multiple domains and typically measured using global cognitive batteries, clinicians and researchers should expect the greater generalizability from multidomain CCT. Several methods for adapting training content to individual cognitive profiles have been developed and are currently undergoing clinical trials, especially within multicomponent dementia prevention trials.^24^ What methods can increase the efficacy of CCT and, importantly, whether these can slow down cognitive decline remains to be investigated in future trials and more specific meta-analysis methods.

### Limitations

To the best of our knowledge, this is the largest systematic review of CCT in older adults to date, and the first to perform a network meta-analysis. Since trials tend to report multiple outcome measures, we used RVE analyses to account for non-independence of effect sizes within studies. Whereas this is an efficient approach that allowed us to detect more heterogeneity and increase the power of meta-regression analyses, a limitation of RVE is that it allows to model dependence due to nesting of multiple outcome measures (correlational model) or groups within studies (hierarchal model), but not both. We used a correlational model as nearly all studies reported multiple outcome measures while only 23 reported more than two eligible arms. Consequently, estimations of weights and heterogeneity did not account for possible differences between subgroups, which may have affected the efficiency of the model specifications.^14^ A method for combining the two working models has been very recently proposed^25^ and may counter this problem in future meta-analyses.

Accounting for dependency in a network meta-analysis is even more challenging,^26^ and we are not aware of viable solutions for generating arm-level composites at this stage. To limit the effect of this problem on our estimates we combined effect sizes within arms into a single estimate using the same correlation-based method^27^ used in our previous univariate meta-analysis of CCT.^5^ Despite applying a large coefficient (r=0.8, as in the RVE model) for estimated correlated variance within arms, there was no detectable heterogeneity in the full network, limiting the precision of our estimates. Nevertheless, imprecision of the main analysis was not substantially greater than the RVE estimates, and arguably less prone to bias compared to selection of outcome measures.

Finally, we did not include data beyond post-training assessments for two main reasons. First, including such data will introduce selection bias as the majority of studies implemented relatively short training periods and did not report long-term outcomes, with considerable variation in the number and length of follow-ups. Second, while some residual gains can still be apparent several months and perhaps up to a year after a course of CCT, effects are expected to wane over time without further training.^28,29^ There is therefore limited value in testing hypotheses related to effect maintenance as the null is the most likely outcome, especially as effects are measured further away from training cessation. Given the ultimate goal of CCT is to delay cognitive decline, implementing and optimizing long-term booster schedules is the key to maintain CCT effects and should be prioritized over more conventional trial designs that focus on modelling the gradual waning of cognitive gains.

## CONCLUSIONS

CCT is efficacious for overall and domain-specific cognitive performance in healthy older adults, but effect vary across key intervention design factors. Greater efficacy should be expected from multidomain CCT, applied up to 3 times per week, and provided in supervised settings. Future trials should avoid inert control conditions whenever possible and focus on optimizing training protocol, specifically in home settings. Research synthesis efforts can move away from investigating mere efficacy and focus on detecting more specific intervention components and individual predictors of training response.

## Data Availability

All underlying data are available from the corresponding author upon request.

## REFERENCES

1. Deary IJ, Corley J, Gow AJ, et al. Age-associated cognitive decline. Br Med Bull. 2009;92:135–152.

2. National Academies of Sciences Engineering and Medicine. Preventing Cognitive Decline and Dementia: A Way Forward. Washington (DC)2017.

3. World Health Organization. Risk reduction of cognitive decline and dementia: WHO Guidelines. Geneva: World Health Organization;2019.

4. Gavelin HM, Lampit A, Hallock H, Sabates J, Bahar-Fuchs A. Cognition-Oriented Treatments for Older Adults: a Systematic Overview of Systematic Reviews. Neuropsychol Rev. 2020.

5. Lampit A, Hallock H, Valenzuela M. Computerized cognitive training in cognitively healthy older adults: a systematic review and meta-analysis of effect modifiers. PLoS Med. 2014;11(11):e1001756.

6. Simons DJ, Boot WR, Charness N, et al. Do “Brain-Training” Programs Work? Psychol Sci Public Interest. 2016;17(3):103–186.

7. Liberati A, Altman DG, Tetzlaff J, et al. The PRISMA statement for reporting systematic reviews and meta-analyses of studies that evaluate health care interventions: explanation and elaboration. PLoS Med. 2009;6(7):e1000100.

8. Hutton B, Salanti G, Caldwell DM, et al. The PRISMA extension statement for reporting of systematic reviews incorporating network meta-analyses of health care interventions: checklist and explanations. Ann Intern Med. 2015;162(11):777–784.

9. Webb SL, Loh V, Lampit A, Bateman JE, Birney DP. Meta-Analysis of the Effects of Computerized Cognitive Training on Executive Functions: a Cross-Disciplinary Taxonomy for Classifying Outcome Cognitive Factors. Neuropsychol Rev. 2018;28(2):232–250.

10. Sterne JAC, Savovic J, Page MJ, et al. RoB 2: a revised tool for assessing risk of bias in randomised trials. BMJ. 2019;366:4898.

11. Fisher Z, Tipton E, Zhipeng H. robumeta : Robust variance meta_regression. R package version 20. 2017.

12. Pustejovsky J. clubSandwich: Cluster-robust (sandwich) variance estimators with small-sample corrections. 2020.

13. Rücker G, Krahn U, König J, Efthimiou O, Schwarzer G. netmeta: Network meta-analysis using frequentist methods. 2020.

14. Hedges LV, Tipton E, Johnson MC. Robust variance estimation in meta-regression with dependent effect size estimates. Res Synth Methods. 2010;1(1):39–65.

15. Borenstein M, Higgins JP, Hedges LV, Rothstein HR. Basics of meta-analysis: I(2) is not an absolute measure of heterogeneity. Res Synth Methods. 2017;8(1):5–18.

16. Riley RD, Higgins JP, Deeks JJ. Interpretation of random effects meta-analyses. BMJ. 2011;342:d549.

17. Sterne JA, Sutton AJ, Ioannidis JP, et al. Recommendations for examining and interpreting funnel plot asymmetry in meta-analyses of randomised controlled trials. BMJ. 2011;343:d4002.

18. Egger M, Davey Smith G, Schneider M, Minder C. Bias in meta-analysis detected by a simple, graphical test. BMJ. 1997;315(7109):629–634.

19. Rucker G, Schwarzer G. Ranking treatments in frequentist network meta-analysis works without resampling methods. BMC Med Res Methodol. 2015;15:58.

20. Gold SM, Enck P, Hasselmann H, et al. Control conditions for randomised trials of behavioural interventions in psychiatry: a decision framework. Lancet Psychiatry. 2017;4(9):725–732.

21. Nguyen L, Murphy K, Andrews G. Immediate and long-term efficacy of executive functions cognitive training in older adults: A systematic review and meta-analysis. Psychol Bull. 2019;145(7):698–733.

22. Teixeira-Santos AC, Moreira CS, Magalhaes R, et al. Reviewing working memory training gains in healthy older adults: A meta-analytic review of transfer for cognitive outcomes. Neurosci Biobehav Rev. 2019;103:163–177.

23. Jaklevic MC. Watch Your Medicine: Video Game Therapy for Children With ADHD. JAMA. 2020;324(3):224.

24. Kivipelto M, Mangialasche F, Snyder HM, et al. World-Wide FINGERS Network: A global approach to risk reduction and prevention of dementia. Alzheimer’s & dementia : the journal of the Alzheimer’s Association. 2020;16(7):1078–1094.

25. Pustejovsky JE, Tipton E. Meta-Analysis with Robust Variance Estimation: Expanding the Range of Working Models. OSF. osf.io/x8yre. Published 2020. Accessed September 2020.

26. Achana FA, Cooper NJ, Bujkiewicz S, et al. Network meta-analysis of multiple outcome measures accounting for borrowing of information across outcomes. BMC Med Res Methodol. 2014;14:92.

27. Gleser LJ, Olkin I. Stochastically dependent effect sizes. In: Cooper H, Hedges L, Valentine J, eds. The handbook of research synthesis and meta-analysis, 2nd Edition. New York: Russell Sage Foundation; 2009:357–376.

28. Zelinski EM, Spina LM, Yaffe K, et al. Improvement in memory with plasticity-based adaptive cognitive training: results of the 3-month follow-up. J Am Geriatr Soc. 2011;59(2):258–265.

29. Lampit A, Hallock H, Moss R, et al. The timecourse of global cognitive gains from supervised computer-assisted cognitive training: A randomised, active-controlled trial in elderly with multiple dementia risk factors. J Prev Alz Dis. 2014;1(1):33–39.

30. Ackerman PL, Kanfer R, Calderwood C. Use it or lose it? Wii brain exercise practice and reading for domain knowledge. Psychol Aging. 2010;25(4):753–766.

31. Anderson S, White-Schwoch T, Parbery-Clark A, Kraus N. Reversal of age-related neural timing delays with training. Proc Natl Acad Sci U S A. 2013;110(11):4357–4362.

32. Anguera JA, Boccanfuso J, Rintoul JL, et al. Video game training enhances cognitive control in older adults. Nature. 2013;501(7465):97–101.

33. Ball K, Berch DB, Helmers KF, et al. Effects of cognitive training interventions with older adults: a randomized controlled trial. JAMA : the journal of the American Medical Association. 2002;288(18):2271–2281.

34. Ballesteros S, Prieto A, Mayas J, et al. Brain training with non-action video games enhances aspects of cognition in older adults: a randomized controlled trial. Front Aging Neurosci. 2014;6:277.

35. Ballesteros S, Mayas J, Prieto A, Ruiz-Marquez E, Toril P, Reales JM. Effects of Video Game Training on Measures of Selective Attention and Working Memory in Older Adults: Results from a Randomized Controlled Trial. Front Aging Neurosci. 2017;9:354.

36. Barban F, Annicchiarico R, Pantelopoulos S, et al. Protecting cognition from aging and Alzheimer’s disease: a computerized cognitive training combined with reminiscence therapy. Int J Geriatr Psychiatry. 2016;31(4):340–348.

37. Barban F, Annicchiarico R, Melideo M, et al. Reducing Fall Risk with Combined Motor and Cognitive Training in Elderly Fallers. Brain Sci. 2017;7(2).

38. Barnes DE, Santos-Modesitt W, Poelke G, et al. The Mental Activity and eXercise (MAX) trial: a randomized controlled trial to enhance cognitive function in older adults. JAMA Intern Med. 2013;173(9):797–804.

39. Basak C, Boot WR, Voss MW, Kramer AF. Can training in a real-time strategy video game attenuate cognitive decline in older adults? Psychol Aging. 2008;23(4):765–777.

40. Belchior P, Marsiske M, Sisco SM, et al. Video game training to improve selective visual attention in older adults. Comput Human Behav. 2013;29(4):1318–1324.

41. Belchior P, Yam A, Thomas KR, et al. Computer and Videogame Interventions for Older Adults’ Cognitive and Everyday Functioning. Games Health J. 2019;8(2):129–143.

42. Berry AS, Zanto TP, Clapp WC, et al. The influence of perceptual training on working memory in older adults. PLoS One. 2010;5(7):e11537.

43. Boot WR, Champion M, Blakely DP, Wright T, Souders DJ, Charness N. Video games as a means to reduce age-related cognitive decline: attitudes, compliance, and effectiveness. Front Psychol. 2013;4:31.

44. Bottiroli S, Cavallini E. Can computer familiarity regulate the benefits of computer-based memory training in normal aging? A study with an Italian sample of older adults. Neuropsychol Dev Cogn B Aging Neuropsychol Cogn. 2009;16(4):401–418.

45. Bozoki A, Radovanovic M, Winn B, Heeter C, Anthony JC. Effects of a computer-based cognitive exercise program on age-related cognitive decline. Arch Gerontol Geriatr. 2013;57(1):1–7.

46. Brehmer Y, Westerberg H, Backman L. Working-memory training in younger and older adults: training gains, transfer, and maintenance. Front Hum Neurosci. 2012;6:63.

47. Buitenweg JIV, van de Ven RM, Prinssen S, Murre JMJ, Ridderinkhof KR. Cognitive Flexibility Training: A Large-Scale Multimodal Adaptive Active-Control Intervention Study in Healthy Older Adults. Front Hum Neurosci. 2017;11:529.

48. Burki CN, Ludwig C, Chicherio C, de Ribaupierre A. Individual differences in cognitive plasticity: an investigation of training curves in younger and older adults. Psychol Res. 2014.

49. Casutt G, Theill N, Martin M, Keller M, Jancke L. The drive-wise project: Driving simulator training increases real driving performance in healthy older drivers. Front Aging Neurosci. 2014;6(MAY).

50. Chan JS, Wu Q, Liang D, Yan JH. Visuospatial working memory training facilitates visually-aided explicit sequence learning. Acta Psychol (Amst). 2015;161:145–153.

51. Colzato LS, van Muijden J, Band GP, Hommel B. Genetic Modulation of Training and Transfer in Older Adults: BDNF ValMet Polymorphism is Associated with Wider Useful Field of View. Front Psychol. 2011;2:199.

52. Dahlin E, Nyberg L, Backman L, Neely AS. Plasticity of executive functioning in young and older adults: immediate training gains, transfer, and long-term maintenance. Psychol Aging. 2008;23(4):720–730.

53. Desjardins-Crepeau L, Berryman N, Fraser SA, et al. Effects of combined physical and cognitive training on fitness and neuropsychological outcomes in healthy older adults. Clin Interv Aging. 2016;11:1287–1299.

54. Du X, Ji Y, Chen T, Tang Y, Han B. Can working memory capacity be expanded by boosting working memory updating efficiency in older adults? Psychol Aging. 2018;33(8):.

55. Dustman RE, Emmerson RY, Steinhaus LA, Shearer DE, Dustman TJ. The effects of videogame playing on neuropsychological performance of elderly individuals. J Gerontol. 1992;47(3):P168–171.

56. Edwards JD, Wadley VG, Myers RS, Roenker DL, Cissell GM, Ball KK. Transfer of a speed of processing intervention to near and far cognitive functions. Gerontology. 2002;48(5):329–340.

57. Edwards JD, Wadley VG, Vance DE, Wood K, Roenker DL, Ball KK. The impact of speed of processing training on cognitive and everyday performance. Aging Ment Health. 2005;9(3):262–271.

58. Edwards JD, Valdes EG, Peronto C, et al. The Efficacy of InSight Cognitive Training to Improve Useful Field of View Performance: A Brief Report. J Gerontol B Psychol Sci Soc Sci. 2015;70(3):417–422.

59. Eggenberger P, Schumacher V, Angst M, Theill N, de Bruin ED. Does multicomponent physical exercise with simultaneous cognitive training boost cognitive performance in older adults? A 6-month randomized controlled trial with a 1-year follow-up. Clin Interv Aging. 2015;10:1335–1349.

60. Frankenmolen NL, Overdorp EJ, Fasotti L, Claassen J, Kessels RPC, Oosterman JM. Memory Strategy Training in Older Adults with Subjective Memory Complaints: A Randomized Controlled Trial. J Int Neuropsychol Soc. 2018;24(10):1110–1120.

61. Garcia-Campuzano MT, Virues-Ortega J, Smith S, Moussavi Z. Effect of cognitive training targeting associative memory in the elderly: A small randomized trial and a longitudinal evaluation. J Am Geriatr Soc. 2013;61(12):2252–2254.

62. Goghari VM, Lawlor-Savage L. Comparison of Cognitive Change after Working Memory Training and Logic and Planning Training in Healthy Older Adults. Front Aging Neurosci. 2017;9:39.

63. Goldstein J, Cajko L, Oosterbroek M, Michielsen M, Van Houten O, Salverda F. Video Games and the Elderly. Social Behavior and Personality: an international journal. 1997;25(4):345–352.

64. Gronholm-Nyman P, Soveri A, Rinne JO, et al. Limited Effects of Set Shifting Training in Healthy Older Adults. Front Aging Neurosci. 2017;9:69.

65. Guye S, von Bastian CC. Working memory training in older adults: Bayesian evidence supporting the absence of transfer. Psychol Aging. 2017;32(8):732–746.

66. Hynes SM. Internet, home-based cognitive and strategy training with older adults: a study to assess gains to daily life. Aging Clin Exp Res. 2016;28(5):1003–1008.

67. Jaeggi SM, Buschkuehl M, Parlett-Pelleriti CM, et al. Investigating the Effects of Spacing on Working Memory Training Outcome: A Randomized, Controlled, Multisite Trial in Older Adults. J Gerontol B Psychol Sci Soc Sci. 2020;75(6):1181–1192.

68. Ji Y, Wang J, Chen T, Du X, Zhan Y. Plasticity of inhibitory processes and associated far-transfer effects in older adults. Psychol Aging. 2016;31(5):415–429.

69. Kuhn S, Lorenz RC, Weichenberger M, et al. Taking control! Structural and behavioural plasticity in response to game-based inhibition training in older adults. Neuroimage. 2017;156:199–206.

70. Lampit A, Hallock H, Moss R, et al. The Timecourse of Global Cognitive Gains from Supervised Computer-Assisted Cognitive Training: A Randomised, Active-Controlled Trial in Elderly with Multiple Dementia Risk Factors. J Prev Alzheimers Dis. 2014;1(1):33–39.

71. Lange S, Süß H-M. Experimental Evaluation of Near- and Far-Transfer Effects of an Adaptive Multicomponent Working Memory Training. Appl Cogn Psychol. 2015;29(4):502–514.

72. Lee HK, Kent JD, Wendel C, et al. Home-Based, Adaptive Cognitive Training for Cognitively Normal Older adults: Initial Efficacy Trial. J Gerontol B Psychol Sci Soc Sci. 2020;75(6):1144–1154.

73. Legault C, Jennings JM, Katula JA, et al. Designing clinical trials for assessing the effects of cognitive training and physical activity interventions on cognitive outcomes: the Seniors Health and Activity Research Program Pilot (SHARP-P) study, a randomized controlled trial. BMC Geriatr. 2011;11:27.

74. Li KZ, Roudaia E, Lussier M, Bherer L, Leroux A, McKinley PA. Benefits of cognitive dualtask training on balance performance in healthy older adults. J Gerontol A Biol Sci Med Sci. 2010;65(12):1344–1352.

75. Mahncke HW, Connor BB, Appelman J, et al. Memory enhancement in healthy older adults using a brain plasticity-based training program: a randomized, controlled study. Proc Natl Acad Sci U S A. 2006;103(33):12523–12528.

76. Maillot P, Perrot A, Hartley A. Effects of interactive physical-activity video-game training on physical and cognitive function in older adults. Psychol Aging. 2012;27(3):589–600.

77. McAvinue LP, Golemme M, Castorina M, et al. An evaluation of a working memory training scheme in older adults. Front Aging Neurosci. 2013;5:20.

78. Millan-Calenti JC, Lorenzo T, Nunez-Naveira L, Bujan A, Rodriguez-Villamil JL, Maseda A. Efficacy of a computerized cognitive training application on cognition and depressive symptomatology in a group of healthy older adults: A randomized controlled trial. Arch Gerontol Geriatr. 2015;61(3):337–343.

79. Miller KJ, Dye RV, Kim J, et al. Effect of a computerized brain exercise program on cognitive performance in older adults. Am J Geriatr Psychiatry. 2013;21(7):655–663.

80. Mishra J, de Villers-Sidani E, Merzenich M, Gazzaley A. Adaptive training diminishes distractibility in aging across species. Neuron. 2014;84(5):1091–1103.

81. Nilsson J, Lebedev AV, Rydstrom A, Lovden M. Direct-Current Stimulation Does Little to Improve the Outcome of Working Memory Training in Older Adults. Psychol Sci. 2017;28(7):907–920.

82. Nouchi R, Taki Y, Takeuchi H, et al. Brain training game improves executive functions and processing speed in the elderly: a randomized controlled trial. PLoS One. 2012;7(1):e29676.

83. Nouchi R, Saito T, Nouchi H, Kawashima R. Small Acute Benefits of 4 Weeks Processing Speed Training Games on Processing Speed and Inhibition Performance and Depressive Mood in the Healthy Elderly People: Evidence from a Randomized Control Trial. Front Aging Neurosci. 2016;8:302.

84. Nouchi R, Kobayashi A, Nouchi H, Kawashima R. Newly Developed TV-Based Cognitive Training Games Improve Car Driving Skills, Cognitive Functions, and Mood in Healthy Older Adults: Evidence From a Randomized Controlled Trial. Front Aging Neurosci. 2019;11:99.

85. Nozawa T, Taki Y, Kanno A, et al. Effects of Different Types of Cognitive Training on Cognitive Function, Brain Structure, and Driving Safety in Senior Daily Drivers: A Pilot Study. Behav Neurol. 2015;2015:525901.

86. O’Brien JL, Edwards JD, Maxfield ND, Peronto CL, Williams VA, Lister JJ. Cognitive training and selective attention in the aging brain: An electrophysiological study. Clin Neurophysiol. 2013;124(11):2198–2208.

87. Payne BR, Stine-Morrow EAL. The Effects of Home-Based Cognitive Training on Verbal Working Memory and Language Comprehension in Older Adulthood. Front Aging Neurosci. 2017;9:256.

88. Peng H, Wen J, Wang D, Gao Y. The impact of processing speed training on working memory in old adults. Journal of Adult Development. 2012;19(3):150–157.

89. Pereira-Morales AJ, Cruz-Salinas AF, Aponte J, Pereira-Manrique F. Efficacy of a computer-based cognitive training program in older people with subjective memory complaints: a randomized study. Int J Neurosci. 2018;128(1):1–9.

90. Peretz C, Korczyn AD, Shatil E, Aharonson V, Birnboim S, Giladi N. Computer-based, personalized cognitive training versus classical computer games: A randomized double-blind prospective trial of cognitive stimulation. Neuroepidemiology. 2011;36(2):91–99.

91. Pergher V, Wittevrongel B, Tournoy J, Schoenmakers B, Van Hulle MM. N-back training and transfer effects revealed by behavioral responses and EEG. Brain Behav. 2018;8(11):e01136.

92. Perrot A, Maillot P, Hartley A. Cognitive Training Game Versus Action Videogame: Effects on Cognitive Functions in Older Adults. Games Health J. 2019;8(1):35–40.

93. Rasmusson DX, Rebok GW, Bylsma FW, Brandt J. Effects of three types of memory training in normal elderly. Aging, Neuropsychology, and Cognition. 1999;6(1):56–66.

94. Richmond LL, Morrison AB, Chein JM, Olson IR. Working memory training and transfer in older adults. Psychol Aging. 2011;26(4):813–822.

95. Rolle CE, Anguera JA, Skinner SN, Voytek B, Gazzaley A. Enhancing Spatial Attention and Working Memory in Younger and Older Adults. J Cogn Neurosci. 2017;29(9):1483–1497.

96. Salminen T, Frensch P, Strobach T, Schubert T. Age-specific differences of dual n-back training. Neuropsychol Dev Cogn B Aging Neuropsychol Cogn. 2016;23(1):18–39.

97. Sandberg P, Ronnlund M, Nyberg L, Stigsdotter Neely A. Executive process training in young and old adults. Neuropsychol Dev Cogn B Aging Neuropsychol Cogn. 2014;21(5):577–605.

98. Shatil E. Does combined cognitive training and physical activity training enhance cognitive abilities more than either alone? A four-condition randomized controlled trial among healthy older adults. Front Aging Neurosci. 2013;5:8.

99. Shatil E, Mikulecka J, Bellotti F, Bures V. Novel television-based cognitive training improves working memory and executive function. PLoS One. 2014;9(7):e101472.

100. Simon SS, Tusch ES, Feng NC, Hakansson K, Mohammed AH, Daffner KR. Is Computerized Working Memory Training Effective in Healthy Older Adults? Evidence from a Multi-Site, Randomized Controlled Trial. J Alzheimers Dis. 2018;65(3):931–949.

101. Simpson T, Camfield D, Pipingas A, Macpherson H, Stough C. Improved processing speed: Online computer-based cognitive training in older adults. Educational Gerontology. 2012;38(7):445–458.

102. Smith GE, Housen P, Yaffe K, et al. A cognitive training program based on principles of brain plasticity: results from the Improvement in Memory with Plasticity-based Adaptive Cognitive Training (IMPACT) study. J Am Geriatr Soc. 2009;57(4):594–603.

103. Sosa GW, Lagana L. The effects of video game training on the cognitive functioning of older adults: A community-based randomized controlled trial. Arch Gerontol Geriatr. 2019;80:20–30.

104. Souders DJ, Boot WR, Blocker K, Vitale T, Roque NA, Charness N. Evidence for Narrow Transfer after Short-Term Cognitive Training in Older Adults. Front Aging Neurosci. 2017;9:41.

105. Stern Y, Blumen HM, Rich LW, Richards A, Herzberg G, Gopher D. Space Fortress game training and executive control in older adults: a pilot intervention. Neuropsychol Dev Cogn B Aging Neuropsychol Cogn. 2011;18(6):653–677.

106. Ten Brinke LF, Best JR, Chan JLC, et al. The Effects of Computerized Cognitive Training With and Without Physical Exercise on Cognitive Function in Older Adults: An 8-Week Randomized Controlled Trial. J Gerontol A Biol Sci Med Sci. 2020;75(4):755–763.

107. Toril P, Reales JM, Mayas J, Ballesteros S. Video Game Training Enhances Visuospatial Working Memory and Episodic Memory in Older Adults. Front Hum Neurosci. 2016;10:206.

108. van het Reve E, de Bruin ED. Strength-balance supplemented with computerized cognitive training to improve dual task gait and divided attention in older adults: a multicenter randomized-controlled trial. BMC Geriatr. 2014;14:134.

109. van Muijden J, Band GP, Hommel B. Online games training aging brains: limited transfer to cognitive control functions. Front Hum Neurosci. 2012;6:221.

110. Van Vleet TM, DeGutis JM, Merzenich MM, Simpson GV, Zomet A, Dabit S. Targeting alertness to improve cognition in older adults: A preliminary report of benefits in executive function and skill acquisition. Cortex. 2016;82:100–118.

111. Vance D, Dawson J, Wadley V, et al. The accelerate study: The longitudinal effect of speed of processing training on cognitive performance of older adults. Rehabil Psychol. 2007;52(1):89–96.

112. von Bastian CC, Langer N, Jancke L, Oberauer K. Effects of working memory training in young and old adults. Mem Cognit. 2013;41(4):611–624.

113. Wang MY, Chang CY, Su SY. What’s Cooking? - Cognitive Training of Executive Function in the Elderly. Front Psychol. 2011;2:228.

114. Wayne RV, Hamilton C, Jones Huyck J, Johnsrude IS. Working Memory Training and Speech in Noise Comprehension in Older Adults. Front Aging Neurosci. 2016;8:49.

115. Weicker J, Hudl N, Frisch S, et al. WOME: Theory-Based Working Memory Training - A Placebo-Controlled, Double-Blind Evaluation in Older Adults. Front Aging Neurosci. 2018;10:247.

116. West RK, Rabin LA, Silverman JM, Moshier E, Sano M, Beeri MS. Short-term computerized cognitive training does not improve cognition compared to an active control in non-demented adults aged 80 years and above. Int Psychogeriatr. 2020;32(1):65–73.

117. Wolinsky FD, Vander Weg MW, Howren MB, et al. Interim analyses from a randomised controlled trial to improve visual processing speed in older adults: the Iowa Healthy and Active Minds Study. BMJ open. 2011;1(2):e000225.

118. Zimmermann K, von Bastian CC, Rocke C, Martin M, Eschen A. Transfer after process-based object-location memory training in healthy older adults. Psychol Aging. 2016;31(7):798–814.

